# Effect of ginger supplementation on the fecal microbiome in subjects with prior colorectal adenoma

**DOI:** 10.1101/2023.10.04.23295545

**Authors:** Ajay Prakash, Nathan Rubin, Christopher Staley, Guillaume Onyeaghala, Ya-Feng Wen, Aasma Shaukat, Ginger Milne, Robert J. Straka, Timothy R. Church, Anna Prizment

**Affiliations:** Division of Hematology, Oncology, and Transplantation, University of Minnesota Medical School, Minneapolis, Minnesota, United States of America; Biostatistics Core, Masonic Cancer Center, University of Minnesota, Minneapolis, Minnesota, United States of America; Department of Surgery, University of Minnesota Medical School, Minneapolis, Minnesota, United States of America; Department of Experimental and Clinical Pharmacology, College of Pharmacy, University of Minnesota, Minneapolis, Minnesota, United States of America; NYU Langone Medical Center, New York, New York, United States of America; Vanderbilt University Medical Center, Nashville, Tennessee, United States of America; Division of Environmental Health Sciences, University of Minnesota School of Public Health, Minneapolis, Minnesota, United States of America

**Keywords:** Microbiome, colon cancer, cancer risk, population health, clinical trial

## Abstract

Ginger has been associated with a decreased incidence of colorectal cancer (CRC) through reduction in inflammatory pathways and inhibition of tumor growth. Recent pre-clinical models have implicated changes in the gut microbiome as a possible mediator of the ginger effect on CRC. We hypothesized that, in adults previously diagnosed with a colorectal adenoma, ginger supplementation would alter the fecal microbiome in the direction consistent with its CRC-inhibitory effect. Sixty-eight adults were randomized to take either ginger or placebo daily for 6 weeks, with a 6-week washout and longitudinal stool collection throughout. We performed 16S rRNA sequencing and evaluated changes in overall microbial diversity and the relative abundances of pre-specified CRC-associated taxa using mixed-effects logistic regression. Ginger supplementation showed no significant effect on microbial community structure through alpha or beta diversity. Of 10 pre-specified CRC-associated taxa, there were significant decreases in the relative abundances of the genera *Akkermansia* (p<0.001), *Bacteroides* (p=0.018), and *Ruminococcus* (p=0.013) after 6-week treatment with ginger compared to placebo. Ginger supplementation led to decreased abundances of *Akkermansia* and *Bacteroides*, which suggests that ginger may have an inhibitory effect on CRC-associated taxa. Overall, ginger supplementation appears to have a limited effect on gut microbiome in patients with colorectal adenomas.

## INTRODUCTION

Colorectal cancer (CRC) is a significant public health burden, and remains the third-leading cause of cancer death in the US despite significant progress in screening and treatment over the past several decades [1, 2]. The gut microbiome plays a critical role in CRC carcinogenesis and progression [3-5], with numerous taxa implicated in tumorigenesis, a subset noted to be persistently enriched across studies in CRC [6]. Enrichment of these pro-inflammatory taxa also leads to relative depletion of the abundance of anti-inflammatory, butyrate-producing commensal bacteria (e.g. *Lactobacillus*), further harming the colonic microenvironment. But while there is significant literature characterizing the microbial shifts which occur during carcinogenesis, little is known regarding how and whether these shifts can be mitigated with targeted interventions. Recently, data from our group and others have shown that the use of anti-inflammatory agents, like aspirin, leads to persistent shifts in the gut microbiome of healthy adults [7, 8]. In particular, the relative abundances of the commensal, gut-protective taxa *Akkermansia*, *Prevotella*, and *Ruminococcus* were all significantly increased following treatment with aspirin in heathy adults. These data suggest that aspirin may favorably alter the CRC microbiome. Thus, we sought to answer the question of whether other well-known dietary agents may also lead to beneficial shifts in the gut microbiome.

Ginger, the rhizome of the plant *Zingiber officinale*, is a well-characterized anti-inflammatory herbal supplement, which has important activity in the gastrointestinal tract, including anti-flatulent, anti-emetic, and digestive aide properties [9-11]. Ginger constituents have been shown to have antioxidant and anti-inflammatory properties, with some pre-clinical data demonstrating inhibition of tumor growth [10, 12]. These pharmacologic effects appear to be mediated by exosome-like nanoparticles (ELNs), 50-100nm lipid spheroids which contain bioactive molecules, including proteins, lipids, and nucleic acids. Recent studies have demonstrated that ginger ELNs alter the gut microbiome, specifically through metabolite interactions with the taxa *Lactobacillus rhamnosus* [13]. Despite preliminary clinical trials demonstrating decreases in inflammatory markers for those subjects taking ginger supplementation [14-17], whether or not this supplementation may alter the microbiome and decrease CRC-associated microbial taxa, remains an open question.

To address this question, we conducted a double-blinded, randomized, placebo-controlled trial to evaluate whether ginger supplementation alters the gut microbiome, including CRC-associated microbial taxa, in subjects with recently diagnosed colorectal adenomas. We hypothesized that 6 weeks of ginger supplementation would lead to enrichment of commensal taxa like *Akkermansia* and *Ruminococcus* as was seen with aspirin, along with decreases in CRC-associated taxa like *Bacteroides* and *Fusobacterium*. To test this hypothesis, we randomized 68 participants with recently diagnosed colorectal adenomas to either the ginger or placebo arm and collected stool samples before, during, and after the 6-week intervention. We sought to compare changes in microbiome composition in relative abundances of genera previously identified to decrease due to aspirin administration, and in CRC-associated biomarkers between the arms.

## METHODS

### Study Population

This randomized, placebo-controlled, double-blinded study targeted 100 subjects between 50 and 75 years old who live throughout Minnesota. Since the MNCCTN partners with health care organizations throughout Minnesota, study recruitment was tailored to each site. Medical record data query using ICD codes were used to identify 7625 eligible individuals at University of Minnesota (UMN) sites, and 2841 individuals at non-UMN sites. Invitation letters were sent by mail to 900 individuals from UMN sites, with a further 548 subjects invited from non-UMN sites based on site-specific capacity (Figure 1). Of these individuals, 32 subjects from UMN sites and 245 subjects from non-UMN sites screened by phone interview. Criterion for inclusion was the diagnosis of colorectal adenoma within the last five years. Criteria for exclusion included use of any antiplatelet or anticoagulant medication including aspirin or non-steroidal anti-inflammatory drugs (NSAIDs) in the past 30 days; use of any medications for diabetes or hypertension; laxative use in the past 30 days; oral or IV antibiotic use in the previous 3 months; allergy to ginger; gastrointestinal (GI) cancer or any serious GI condition or surgery within 6 months; any serious active medical (cancer, coronary vascular disease) or neuro-psychiatric illness; BMI ≥ 40 or ≤ 17 kg/m^2^; unexplained change in weight of > 4.5 kg within the past 6 months; or major changes in eating habits within the past 3 months. Of screened participants, 69 subjects were consented across all sites, with 1 subject declining participation after consent. At Visit 1, 68 subjects were randomized to the ginger (N = 33) or placebo (N = 35) arm according to a block randomization protocol. The duration of treatment (6 weeks) was based on a prior trial of aspirin in healthy subjects, in which aspirin taken for 6 weeks altered microbiome composition, with this effect being reversed after a 6-week washout [7].

**Figure 1.**
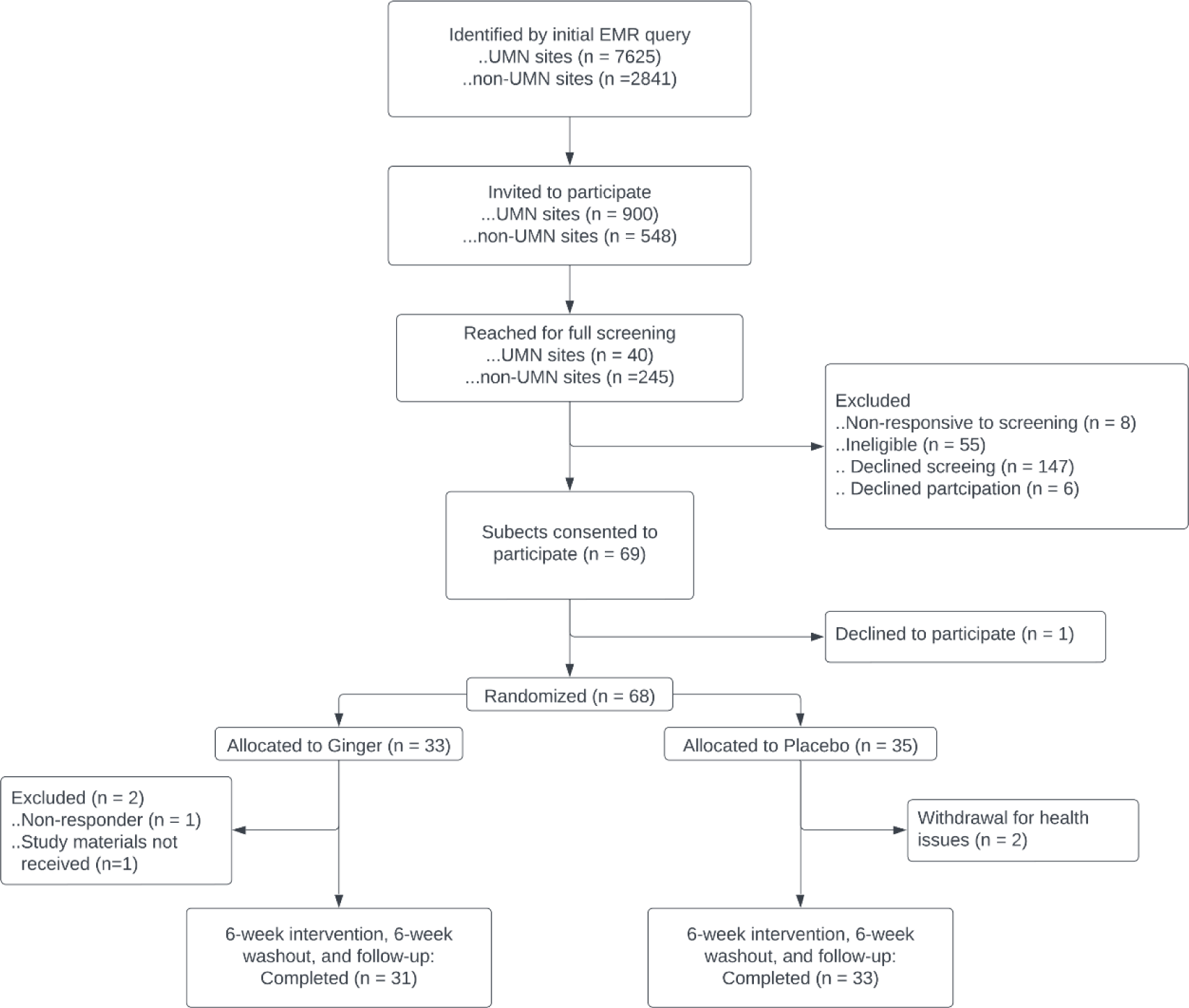
Study Flowchart. Subject population sizes and exclusion criteria for screening, enrollment, and study completion

Additionally, at baseline, subjects were asked a series of questions about health, and medication use. The same questions were repeated after the 6-week treatment/placebo period. Visits were followed up with five phone calls at 3-week intervals (Figure 2) to ask about changes in health status and possible adverse events and to discuss subjects’ upcoming stool collection.

**Figure 2.**
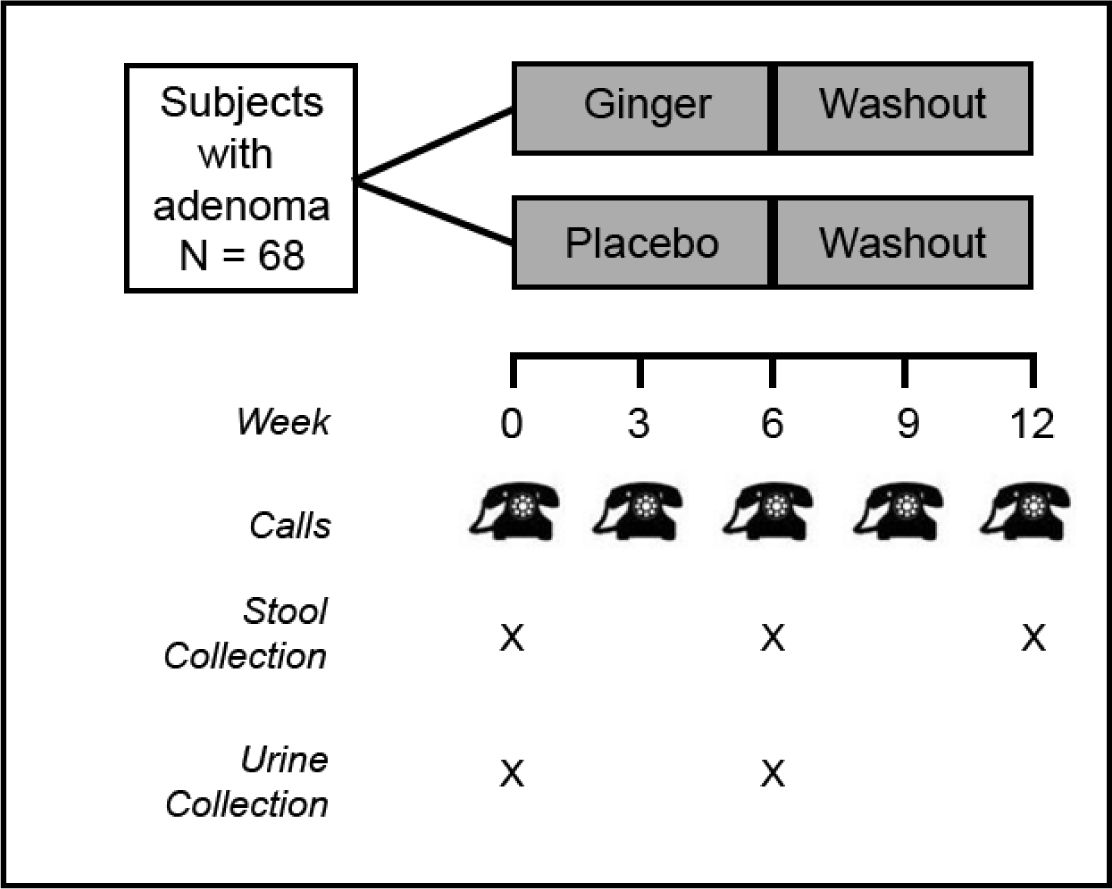
Diagram of interventions and sample collections. Overall study design for ginger treatment and subsequent stool and urine sample collections

### Sample collection and analysis

Urine and stool samples were collected at home by participants as previously described (Figure 1;[7]). Briefly, stool and urine samples were collected in clean, single-use specimen containers. Stool collection kits contained 90% ethanol. Both sample types were placed on ice and transported by secure mail within 72 hours and stored in a -80°C freezer until analysis. Samples from all collections were batch processed. DNA was extracted using the PowerSoil Pro DNA Isolation KIT (MoBio/Qiagen) following manufacturer’s labeling. The V4 hypervariable region of the 16S rRNA gene was amplified and sequenced using the 515F-806R primer set [18], on a MiSeq Illumina platform (2x 300bp paired-end). All sequencing was performed by the University of Minnesota Genomics Center [19]. Negative controls were included and demonstrated no amplification. Sequence data was deposited in the Sequence Read Archive under accession number SRP337967.

Urinary PGE-M levels, an biomarker known to be associated with CRC risk [20], were measured using high-performance liquid chromatography/mass spectrometry (HPLC/MS) and normalized for creatinine, measured using the Enzo Life Sciences test kit (P/N ADI1907030A; ng PGE-M/ mg creatinine). Both PGE-M and urinary creatinine were measured in the Eicosanoid Core Laboratory at Vanderbilt University Medical Center.

### Supplement preparation and treatment compliance

Ginger and placebo (lactose) capsules were compounded by Pure Encapsulations Inc. (a subsidiary of Atrium Innovations Inc.) and packaged by Fairview Investigational Drug Services (IDS), according to the guidelines created by the International Conference on Harmonization of Technical Requirements for Registration of Pharmaceuticals for Human Use (ICH), Good Clinical Practice (GCP) and Good Manufacturing Practice (GMP). Blind-coded study bottles of 60 pills containing either ginger capsules or placebo were prepared for a 6-week daily treatment by block randomization, performed by the University of Minnesota Biostatistics Core. Randomization was implemented by the IDS, with labeling per applicable regulations. Adherence was assessed by comparing the number of capsules returned after treatment completion to the expected number based on treatment duration. Of note, no plant sampling was performed in the study.

### Bioinformatic analysis

Sequence data were processed and analyzed using Mothur ver. 1.41.1 [21, 22]. The number of sequences generated for the analysis, following initial processing, was 7,889,878, and the median number of sequences per sample was 40,395. High-quality sequences were aligned against the SILVA database ver. 138, and chimeric sequences were identified and removed using UCHIME software. Samples were rarefied to 8,800 sequence reads per sample to reduce bias in comparisons. To estimate the proportion of the gut microbiome operational taxonomic units (OTUs) represented in our samples, the mean Good’s coverage among baseline samples was calculated. Clustering of OTUs was performed at 99% identity. Taxonomic classification was performed against the version 18 data release from the Ribosomal Database Project.

### Statistical analysis

Microbiome diversity was evaluated using Mothur. Alpha diversity was calculated as the Shannon and Chao1 indices. Beta diversity was calculated using Bray-Curtis dissimilarity matrices, evaluated using analysis of similarity (ANOSIM) [23], and visualized by ordination using principal coordinates analysis (PCoA) [24]. Longitudinal analysis was performed using the R package SplinectomeR [25].

Mixed-effects logistic regression models with a random intercept for subject were used to evaluate the effect of ginger on the relative abundance of each taxon at week 6. This model specified the outcome measure as taxonomic relative abundance (i.e. proportion), with the covariate terms of visit (week 6 vs. baseline), treatment (ginger vs. control), and the interaction between visit and treatment (week 6 x ginger). This interaction is of primary interest and its’ odds ratios and p-values are presented in the results. The counts of each taxa were also used in each model as a weighting factor. Linear mixed-effects regression models were used to evaluate the effect of ginger on mean PGE-M, again using a random intercept for subject and covariates of visit, treatment, and the treatment × visit interaction. Mixed-effects models were performed using the R package lmerTest [26]. P-values less than 0.05 were considered statistically significant, and no adjustments were made for multiple testing.

### Ethics approval and consent to participate

This clinical trial was reviewed and approved by the University of Minnesota Institutional Review Board (Approval Number: SSU00065591; Approval Date: 7 Sep 2018). It was performed with the express written informed consent of all participants, and in accordance with the relevant guidelines and regulations. Clinicaltrials.gov: NCT03268655; August 31, 2017.

## RESULTS

### Study population characteristics

A total of 68 participants were successfully randomized to either ginger or placebo, with no statistically significant differences in relevant baseline demographic variables, including age, sex, BMI, or adenoma diagnosis year (Table 1). Study retention remained high at all sites, with 65 subjects enrolling out of 68 screened, and 64 subjects completing the study (94% retention).

**Table 1.** Demographics. Demographic characteristics of subjects included within the final study, including subject recruitment location (partner), age, sex, BMI, and year of adenoma diagnosis.

|  | Ginger<br>(N=33) | Placebo<br>(N=35) | Total (N=68) |
| --- | --- | --- | --- |
| **Partner** |  |  |  |
| Essentia | 5 (15.2%) | 5 (14.3%) | 10 (14.7%) |
| Fairview | 14 (42.4%) | 14 (40.0%) | 28 (41.2%) |
| Mayo | 5 (15.2%) | 6 (17.1%) | 11 (16.2%) |
| MMCORC | 1 (3.0%) | 2 (5.7%) | 3 (4.4%) |
| UMN | 8 (24.2%) | 8 (22.9%) | 16 (23.5%) |
| **Age** |  |  |  |
| Mean (SD) | 60.3 (4.9) | 60.1 (6.5) | 60.2 (5.7) |
| Median (Range) | 61.0 (50.0, 68.0) | 61.0 (50.0, 74.0) | 61.0 (50.0, 74.0) |
| **Sex** |  |  |  |
| Female | 24 (72.7%) | 22 (62.9%) | 46 (67.6%) |
| Male | 9 (27.3%) | 13 (37.1%) | 22 (32.4%) |
| **BMI** |  |  |  |
| Mean (SD) | 28.7 (4.8) | 28.3 (4.7) | 28.5 (4.7) |
| Median (Range) | 28.1 (21.2, 39.7) | 28.3 (20.2, 37.8) | 28.2 (20.2, 39.7) |
| **Adenoma Dx Year** |  |  |  |
| 2017 - Present | 27 (81.8%) | 28 (80.0%) | 55 (80.9%) |
| < 2017 | 0 (0.0%) | 2 (5.7%) | 2 (2.9%) |
| Unknown (UMN use only) | 6 (18.2%) | 5 (14.3%) | 11 (16.2%) |

### Intervention adherence

Study intervention adherence was high, as measured by pills returned out of 168. Mean number of capsules taken by study participants was 141.1 (SD = 55.2) and 152.2 (SD = 34.0), respectively in the ginger and placebo groups (Table 2). 90.7% of participants reported at least 70% adherence, with 76.9% reporting 100% adherence. Median adherence rate with for ginger and placebo were 92.2% and 90.1%, respectively.

**Table 2.**
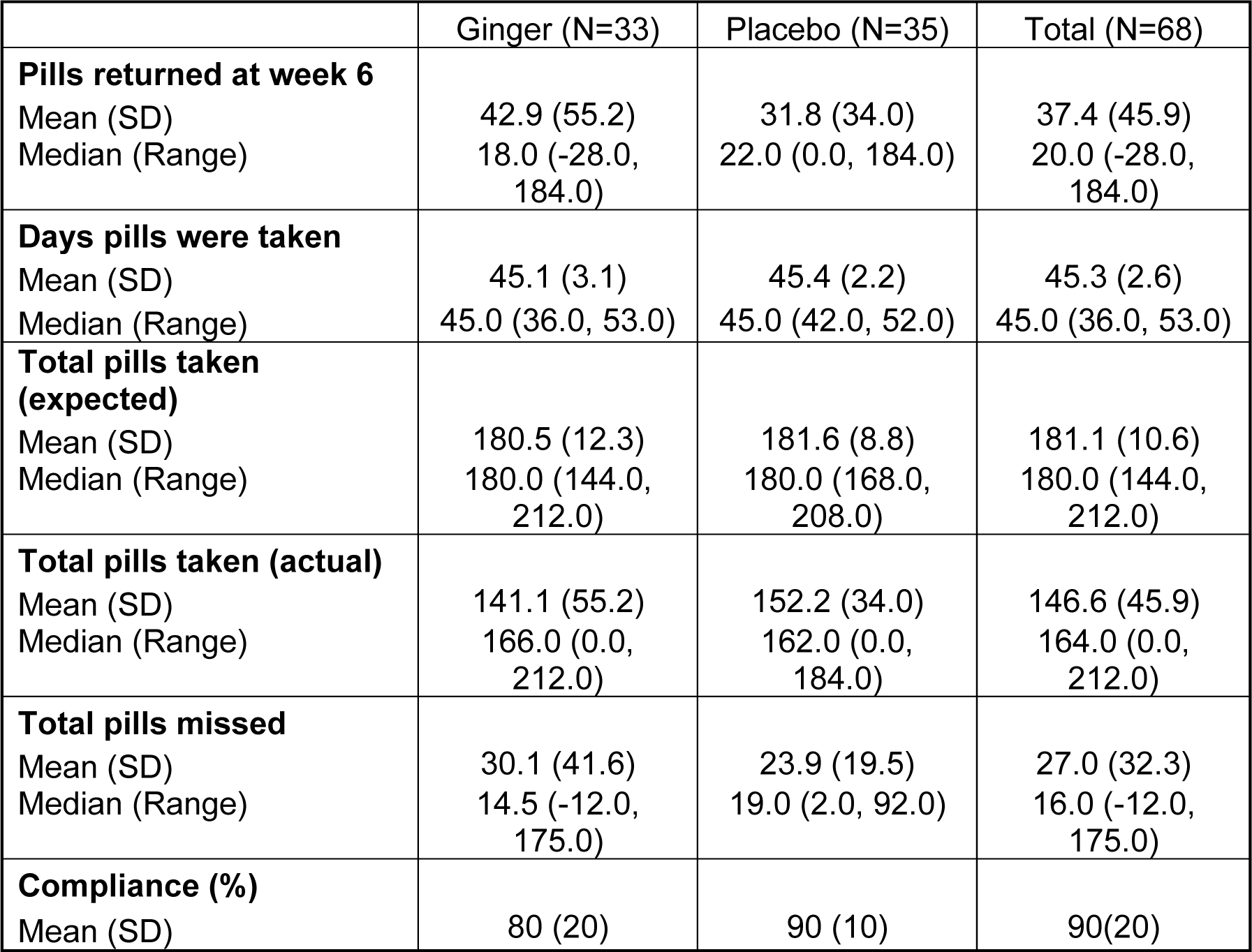
Pill Adherence. Mean and median pills taken of either Ginger or Placebo. Negative value in range of pills missed indicate additional pills which were inadvertently taken

|  | Ginger (N=33) | Placebo (N=35) | Total (N=68) |
| --- | --- | --- | --- |
| **Pills returned at week 6** |  |  |  |
| Mean (SD) | 42.9 (55.2) | 31.8 (34.0) | 37.4 (45.9) |
| Median (Range) | 18.0 (-28.0, 184.0) | 22.0 (0.0, 184.0) | 20.0 (-28.0, 184.0) |
| **Days pills were taken** |  |  |  |
| Mean (SD) | 45.1 (3.1) | 45.4 (2.2) | 45.3 (2.6) |
| Median (Range) | 45.0 (36.0, 53.0) | 45.0 (42.0, 52.0) | 45.0 (36.0, 53.0) |
| **Total pills taken (expected)** |  |  |  |
| Mean (SD) | 180.5 (12.3) | 181.6 (8.8) | 181.1 (10.6) |
| Median (Range) | 180.0 (144.0, 212.0) | 180.0 (168.0, 208.0) | 180.0 (144.0, 212.0) |
| **Total pills taken (actual)** |  |  |  |
| Mean (SD) | 141.1 (55.2) | 152.2 (34.0) | 146.6 (45.9) |
| Median (Range) | 166.0 (0.0, 212.0) | 162.0 (0.0, 184.0) | 164.0 (0.0, 212.0) |
| **Total pills missed** |  |  |  |
| Mean (SD) | 30.1 (41.6) | 23.9 (19.5) | 27.0 (32.3) |
| Median (Range) | 14.5 (-12.0, 175.0) | 19.0 (2.0, 92.0) | 16.0 (-12.0, 175.0) |
| **Compliance (%)** |  |  |  |
| Mean (SD) | 80 (20) | 90 (10) | 90(20) |

### Inflammatory biomarkers

No significant changes were noted in CRC-associated inflammatory markers While change in the mean PGE-M (mPGE-M) levels showed no statistically significant difference following 6 weeks of treatment with either ginger or placebo, mPGE-M levels trended to decrease following ginger treatment. Mean PGE-M levels decreased by 1.29 mg/dL (SD 9.19) in subjects treated with ginger and increased by 0.87 mg/dL (SD 4.88; p = 0.241) in subjects treated with placebo (Supp Table S1). Levels of other cancer-associated biomolecules, like TxB2, did not change significantly over the 6 weeks of treatment with either ginger or placebo.

### Microbial diversity in subjects with treated colorectal adenomas

Most of the bacterial community was captured, with 99.0 ± 0.8% mean estimated Good’s coverage. No significant differences in subject alpha-diversity, as measured by Shannon or Chao1 indices were noted in subjects undergoing treatment with either ginger or placebo (Figure 3; Shannon p = 0.971, and Chao p = 0.431). The microbiome composition from samples collected after treatment (week 6) shifted similarly in both ginger- and placebo-treated subjects (p = 0.800), suggesting that overall microbial shifts were not significantly altered by ginger. This finding was supported by our beta diversity analysis, which showed no significant temporal differences within the ginger and placebo groups when analyzed using Bray-Curtis dissimilarities (ANOSIM R = -0.035 and -0.034, respectively, p = 1.000 for both groups).

**Figure 3.**
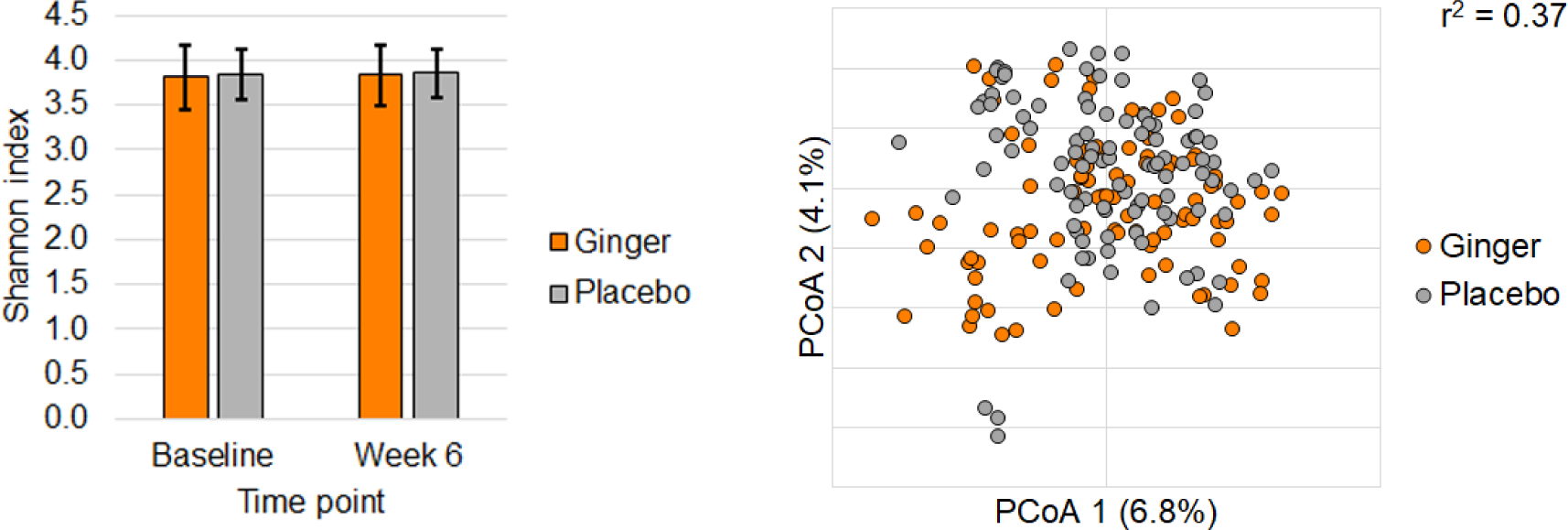
Microbiome structure in ginger supplementation. Alpha and beta Diversity analysis of microbiome composition between serial stool samples in subjects being treated with either ginger (orange) or placebo (grey)

Longitudinal taxonomic analysis by SplinectomeR showed no significant differences in relative abundances of predominant genera between ginger and placebo groups over the 6-week treatment period. While the abundance of *Faecalibacterium* decreased during ginger treatment, a larger decrease occurred following treatment washout, suggesting a possible post-treatment effect of ginger supplementation on *Faecalibacterium* abundance.

### Relative abundance of previously indicated taxa following ginger treatment

Of the pre-specified taxa associated with ginger supplementation in prior publications [11, 14, 17, 27], we noted shifts in the relative abundances of three genera following 6 weeks of ginger treatment (Figure 4). We noted a decrease in the relative abundances of the genera *Bacteroides* (OR 0.95; p = 0.018), *Akkermansia* (OR 0.71; p < 0.001), and *Ruminococcus* (OR 0.84; p = 0.013) in subjects who received ginger supplementation relative to placebo (Figure 3). In the mixed effects analysis, regression coefficients for the interaction term were significant at six weeks of treatment vs baseline for all three genera. There was no significance in the regression coefficients for the genera *Faecalibacterium* (OR 0.94; p = 0.383) and *Prevotella* (OR 0.92; p = 0.306), in contrast to our findings when evaluating the effects of aspirin.

**Figure 4.**
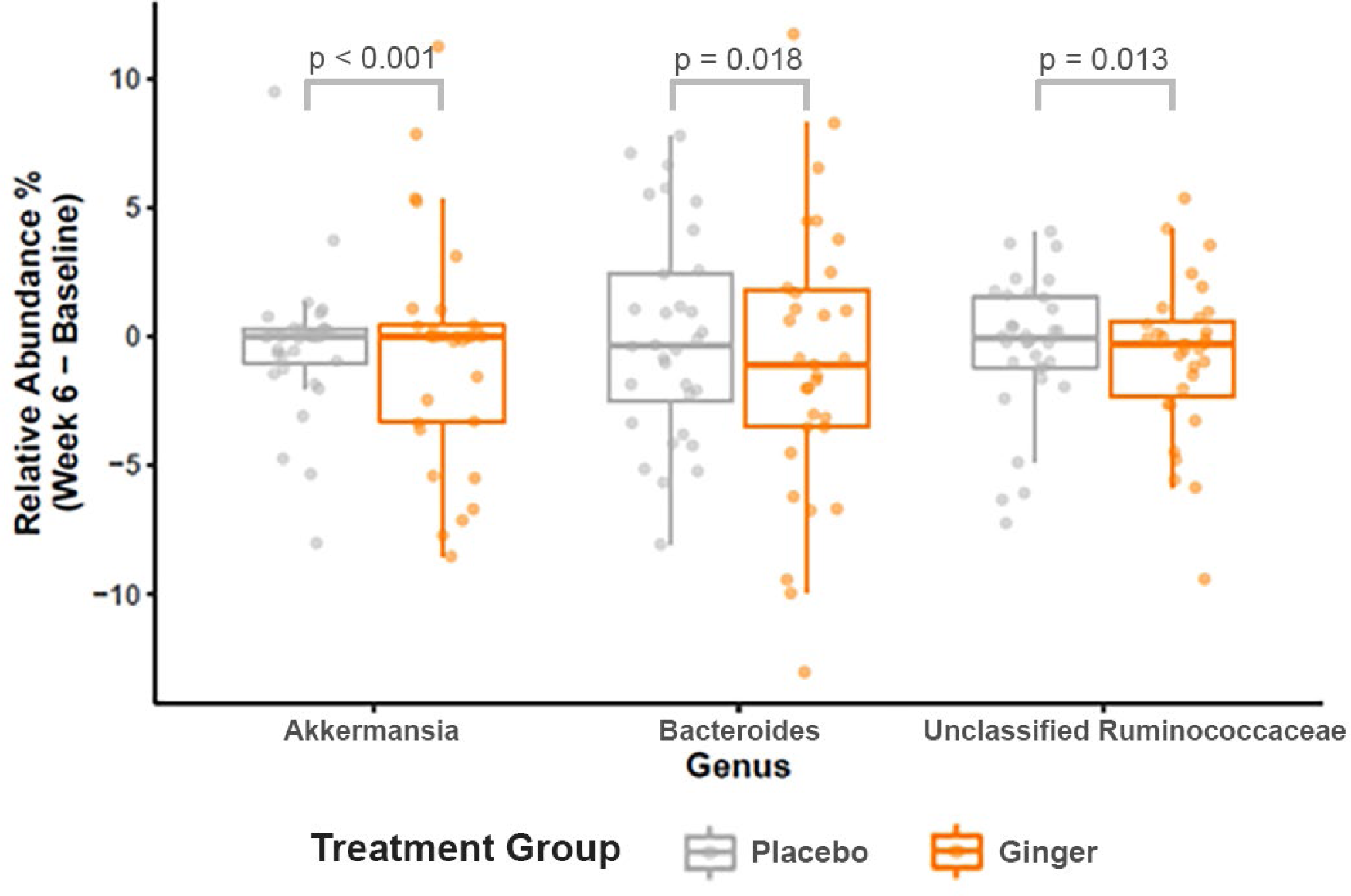
Change in taxonomic mean relative abundance with ginger supplementation. Mean relative abundance and 95% confidence intervals of three CRC-associated taxa which demonstrated significant shifts between ginger (orange) and placebo (grey) during the 6-week intervention

## DISCUSSION

This double-blinded, placebo-controlled, randomized pilot-study of ginger supplementation in subjects with prior colorectal adenoma demonstrates limited changes in the fecal bacterial community between subjects taking ginger and placebo. In particular, the anti-inflammatory genera *Akkermansia*, *Bacteroides*, and *Ruminococcus* exhibited reductions for participants receiving ginger supplementation. However, there were no statistically significant shifts in the overall microbiome as measured by alpha or beta diversity, or in ginger- and inflammatory markers such as PGE-M. These findings suggest that ginger produces a real, but limited change in the fecal microbiome.

Our study did not show enrichment of *Lactobacillus*, as has been demonstrated previously [13]. The most likely source of variance in the effect of ginger on the microbiome is due to fundamental differences in the murine model system used in pre-clinical studies and the human gut being studied here. The immune and microbial architectures fundamentally differ between mouse and human models [28, 29], which may alter the impact of ginger supplementation. In addition, there may exist differential processing of ginger metabolites in the murine model relative to the human gut which may again alter shifts in microbial abundance. These questions require further study to determine if earlier findings regarding the effect of ginger on the microbiome are generalizable to humans. Indeed, early-phase clinical trials in humans appear to support this hypothesis, with relatively muted shifts in inflammatory markers [14].

Our finding of a limited effect of ginger supplementation on the fecal microbiome is biologically plausible and may be related to the differing concentrations of bioactive molecules within different ginger supplements. Earlier studies demonstrating significant shifts in the ginger microbiome utilized a ginger puree with multiple centrifugation steps to isolate supernatant with exosome-like nanoparticles [11, 13]. Prior pilot clinical studies that examined the ginger effect on cell-cycle biomarkers, inflammatory eicosanoids, and PGE levels utilized a compounded form of ginger supplementation which was normalized to gingerol concentration [15-17]. Given that concentrations of bioactive components of ginger may differ between products typically sold within a pharmacy and even differ between lots of the same product, our finding and that of others of a variable effect of ginger on gut inflammation and the microbiome across studies is understandable. Thus, if shifts in the microbiome induced by ginger are of a similar magnitude to those variances in ginger supplementation, they may be difficult to identify using existing methodologies. Further studies may be necessary to identify the specific ginger metabolite(s) and concentration(s) of those components necessary to produce a clear signal in microbial shifts within humans.

A notable strength of our study design and analysis is its size and robust design. Our data provide the first evidence from a randomized clinical trial that ginger induces specific shifts in the fecal microbiome relative to placebo. Inclusion of a placebo-controlled arm reduces the influence of unmeasured confounders, including the normal shift of the gut microbiome during the study period. We generally do not expect these shifts to alter our observations on microbial diversity given the relative stability of the microbiome over the course of months [30, 31]. However, even if these shifts were to be large, their random nature would tend to bias our findings toward Type II error, strengthening our overall conclusions.

A limitation of our study is the relatively brief duration of our intervention, which prevents the identification of more subtle shifts in the microbiome or of shifts in low-abundance, but biologically meaningful, taxa. These include taxa such as *Fusobacterium nucleatum*, which is almost never identified in healthy subjects [3, 4]. Finally, our placebo capsules are primarily constructed of methylcellulose, which is distinct in composition and quantity from the ginger supplements used in prior studies, potentially limiting cross-trial comparison of ginger-specific effects.

In conclusion, our double-blinded, randomized, placebo-controlled pilot trial suggests overall, that ginger supplementation induces limited shifts in the microbiome, with some select taxa demonstrating a distinct reduction. Those taxa associated with a decline were previously shown to be associated with inflammation reduction. Although these are preliminary findings, they may inform considerations of the design of subsequent follow-up studies. These considerations include a larger number of participants in a clinical trial or tighter control of the nature and concentrations of the bioactive components of ginger administered to the supplementation subgroup. Finally, consideration of various combinations of nutritional supplementation (e.g. turmeric, resveratrol) to induce larger effects on inflammation and CRC risk may also be considered.

### Data Availability Statement

The datasets generated and/or analyzed during the current study are available in the Sequence Read Archive under accession number SRP337967.

https://www.ncbi.nlm.nih.gov/sra/?term=SRP337967

## Abbreviations

ANOSIM: Analysis of similarity
CRC: Colorectal cancer
ELN: Exosome-like nanoparticle
GCP: Good Clinical Practice
GMP: Good Manufacturing Practice
GI: Gastrointestinal
ICH: International Council for Harmonisation of Technical Requirements for Pharmaceutials for Human Use
IDS: Investigational drug service
mPGE-M: Mean prostaglandin-E metabolite
NSAID: Non-steroidal anti-inflammatory drug
OTU: Operational taxonomic unit
PCoA: Principle coordinates analysis
UMN: University of Minnesota

## Author Contribution Statement

AjP interpreted data and was a major contributor in writing the manuscript and prepared figures and tables in coordination with co-authors. NR, CS, GO, and YFW analyzed and interpreted data. CS generated figure 3 while NR generated figure 3. GM performed external sample processing and analysis. AS, RJS, TRC, and AnP developed the project, provided associated funding, and assisted in data interpretation. All authors read and approved the final manuscript.

## Additional Information

The authors declare that they have no competing interests

