## Supplemental Tables for "Effect of ginger supplementation on the fecal microbiome in subjects with prior colorectal adenoma"

**Supplemental Table S1. Cancer associated urinary biomolecule levels.** Urinary levels of creatinine, 11-dehydro-thromboxane B2, and prostaglandin E2

### Ginger

| | week 0 (1) (N=31) | week 6 (2) (N=31) | V2 - V1 (N=31) |
|---|---|---|---|
| **creatinine** | | | |
| Mean (SD) | 1.04 (0.58) | 1.18 (0.68) | 0.14 (0.75) |
| Median (Range) | 0.94 (0.11, 2.14) | 0.99 (0.23, 2.83) | 0.08 (-1.42, 2.45) |
| **dehydo_txb2** | | | |
| Mean (SD) | 0.41 (0.16) | 0.40 (0.14) | -0.01 (0.12) |
| Median (Range) | 0.38 (0.21, 0.96) | 0.40 (0.17, 0.84) | 0.01 (-0.34, 0.26) |
| **pge_m** | | | |
| Mean (SD) | 13.05 (10.21) | 11.76 (9.37) | -1.29 (9.19) |
| Median (Range) | 9.06 (2.50, 39.60) | 9.45 (3.50, 43.75) | 0.11 (-28.63, 14.61) |
| **tn_e** | | | |
| Mean (SD) | 2.56 (1.99) | 2.95 (2.27) | 0.39 (0.80) |
| Median (Range) | 2.16 (0.50, 10.28) | 2.48 (0.62, 11.06) | 0.40 (-1.56, 2.32) |

### Placebo

| | week 0 (1) (N=33) | week 6 (2) (N=33) | V2 - V1 (N=33) |
|---|---|---|---|
| **creatinine** | | | |
| Mean (SD) | 1.01 (0.57) | 1.00 (0.52) | -0.01 (0.44) |
| Median (Range) | 0.87 (0.24, 1.91) | 0.93 (0.20, 2.35) | -0.04 (-1.01, 0.82) |
| **dehydo_txb2** | | | |
| Mean (SD) | 0.41 (0.16) | 0.40 (0.15) | 0.00 (0.15) |
| Median (Range) | 0.41 (0.00, 0.77) | 0.39 (0.18, 0.79) | -0.02 (-0.25, 0.44) |
| **pge_m** | | | |
| Mean (SD) | 9.34 (3.76) | 10.22 (4.95) | 0.87 (4.88) |
| Median (Range) | 9.65 (3.20, 18.87) | 10.17 (3.45, 22.44) | 0.88 (-9.06, 12.52) |
| **tn_e** | | | |
| Mean (SD) | 3.40 (3.41) | 3.16 (2.09) | -0.24 (2.17) |
